# Heterogeneity in excess respiratory acute care utilization during wildfire smoke waves in California

**DOI:** 10.64898/2026.11.19.26368512

**Authors:** Yiqun Ma, Arnab K. Dey, Rosana Aguilera, Noemie Letellier, Pierre Scala, Alexander Gershunov, Aron Walker, Caitlin G. Jones-Ngo, Michael Jerrett, Rachel Connolly, Neeta Thakur, Miriam E. Marlier, Tarik Benmarhnia

## Abstract

**Rationale:** Wildfire smoke is an increasing public health threat, yet most epidemiologic studies estimate average smoke-health associations across broad spatial or temporal scales. Such approaches may obscure substantial heterogeneity across individual smoke waves, leaving an important evidence gap for dynamic public health preparedness, risk communication, and clinical practice.

**Objectives:** To estimate event-specific excess respiratory acute care utilization during smoke waves and identify smoke wave features associated with higher excess utilization.

**Methods:** We conducted a statewide study of 18,271,754 respiratory acute care encounters in California from 2006 to 2019. A smoke wave was defined as at least 2 consecutive days during which county-level wildfire smoke fine particulate matter (PM_2.5_) exceeded the 95^th^ percentile over the study period. We applied a machine learning-integrated two-stage interrupted time-series design to estimate excess respiratory acute care utilization during each event. We then used meta-regressions to examine multiple spatiotemporally varying effect modifiers such as smoke wave duration, intensity, timing, and co-occurring climate and meteorological conditions.

**Measurements and Main Results:** A total of 1,249 smoke waves were identified and examined in this study. Excess respiratory acute care utilization varied substantially across smoke waves. Higher excess utilization was associated with longer smoke wave duration, greater intensity, winter occurrence, short recovery period, overlap with holidays, drought conditions, or co-occurrence with precipitation or extreme heat events.

**Conclusions:** This study proposes a novel analytical framework to systematically evaluate spatiotemporal drivers of heterogeneity in wildfire smoke respiratory health effects. The findings can inform dynamic preparedness and public health response to smoke events.

## Introduction

Wildfire smoke is a growing public health concern and is projected to intensify under climate change.^1,2^ A substantial body of literature has linked wildfire smoke exposure to adverse health outcomes, with particularly consistent evidence for acute respiratory outcomes following short-term exposure to smoke fine particulate matter (PM_2.5_).^3–8^ The vast majority of prior studies, however, have analyzed wildfire smoke as a homogeneous exposure and estimated a single average effect for an entire country, region, or state, over multiple years or decades.^3–5,9–11^ Although these average effects are important for quantifying overall burden, such a one-size-fits-all approach may obscure critical spatiotemporal heterogeneity across specific wildfire smoke episodes (commonly referred to as “smoke waves”^2^) and provide limited insight into why some smoke waves have more substantial health impacts than others. Better understanding the drivers of the most impactful wildfire smoke events is essential to inform targeted and dynamic wildfire smoke preparedness, risk communication, resource allocation, and clinical readiness for patients with respiratory disease.^12–15^

Conceptually, heterogeneity in the health impacts of smoke waves may arise from three categories of effect modifiers: smoke wave characteristics (e.g., duration and intensity), time-fixed or slowly varying contextual factors (e.g., geographic location and characteristics of exposed communities), and time-varying factors (e.g., season and co-occurring meteorological conditions).

First, characteristics of the smoke wave itself may plausibly influence its respiratory health impacts. Prior evidence indicates that respiratory disease risks increase with smoke wave intensity.^16,17^ Remarkably, although over 40 % of western U.S. counties are anticipated to have longer smoke waves under climate change,^2^ the role of exposure duration in health impacts remains unclear. This is partly because most prior studies have modeled wildfire smoke as daily air pollutant concentrations (e.g., smoke PM_2.5_) and estimated average exposure-response associations across days rather than modeling discrete multi-day smoke episodes with measurable duration.^18^ In addition, smoke waves can exhibit distinct diurnal exposure patterns (e.g., smoke peaking during the day or at night),^19^ but little is known about whether the diurnal timing of exposure matters for associated health effects.

Second, the epidemiologic literature on heterogeneity in wildfire smoke health impacts has more commonly focused on time-fixed modifying factors (such as socio-demographic or neighborhood characteristics) and spatial variation.^20–22^ These studies have provided important evidence that the respiratory health impacts of wildfire smoke vary considerably across populations and places.

Finally, time-varying factors may also shape the health impacts of wildfire smoke, but evidence remains fragmented. There are mixed findings on how season modifies the effect of PM_2.5_ on short-term respiratory health^23^ and few studies have focused specifically on wildfire smoke. The health consequences of smoke waves may be amplified when they co-occur with other climate-related hazards;^24^ limited but growing evidence suggests synergistic health effects of joint exposure to extreme heat and smoke PM_2.5_.^25–27^ Little is known, however, about whether other compounding climate exposures, such as drought, precipitation, and Santa Ana winds (hot, dry, downslope winds in Southern California), may exacerbate the health impacts of smoke waves. A more comprehensive understanding of the roles of these co-occurring exposures in shaping the relationship between smoke and health is urgently needed, as anthropogenic climate change is likely to increase the burden of such compound events.^24^

The limited understanding of spatiotemporal heterogeneity in smoke-health associations, especially effect modification by time-varying factors, partly reflects methodological challenges. Traditional epidemiologic approaches typically estimate an overall exposure-response association and then assess heterogeneity using stratified analyses or interaction terms.^28^ Although these approaches are useful, they often require pre-specification of a limited set of modifiers and can become unstable or difficult to interpret when multiple correlated factors are considered simultaneously or when data are sparse within some strata. More modern approaches, including spatial Bayesian hierarchical models^20^ and generalized random forests,^21^ have expanded the ability to examine a wide range of time-fixed effect modifiers, but they are not well suited for time-varying effect modifiers. In addition, some modifiers are naturally event-level attributes, making them difficult to evaluate within models that treat smoke as a daily exposure.

Capitalizing on the timing of natural experiments, the recently proposed machine learning (ML)-integrated two-stage interrupted time-series (ITS) design offers a powerful framework for estimating the acute health impacts of events with clearly defined periods.^29,30^ This approach is particularly well suited for smoke waves, which occur over discrete time windows and may trigger short-term changes in respiratory outcomes. Moreover, by estimating the counterfactual disease burden expected in the absence of each event, this approach can directly generate event-specific excess burden estimates.^30^ These event-specific estimates (and related statistical uncertainty) can then be used to evaluate a wide range of event-level attributes as potential effect modifiers, particularly smoke wave characteristics and time-varying factors.

California is among the states most affected by wildfire smoke, and its diverse geographic and climate conditions, as well as its distinct fire regimes (i.e., fuel- and wind-dominated fires),^31^ make it a valuable setting for studying the heterogeneity in health impacts across smoke waves. In this study, we develop a novel analytical framework to estimate event-specific excess respiratory acute care utilization across 1,249 smoke waves in California from 2006 to 2019 and identify smoke wave characteristics and time-varying features associated with higher excess respiratory-related utilization burden.

## Methods

### Respiratory acute care utilization

We obtained data on daily unscheduled inpatient admissions and emergency department visits in California from 2006 to 2019 from the California Department of Health Care Access and Information.^32^ We refer to these encounters collectively as “acute care utilization” or “acute care encounters.” The outcome of interest was respiratory acute care utilization, identified using the primary International Classification of Diseases, Ninth or Tenth Revision codes (ICD-9: 460-519; ICD-10: J00-J99). Individual-level records were then aggregated into county-level daily counts of respiratory acute care encounters based on patients’ county of residence. Among California’s 58 counties, we included the 56 with populations of at least 5,000 in this analysis to improve statistical precision.

### Smoke waves

Daily county-level wildfire smoke PM_2.5_ concentrations in California from 2006 to 2019 were calculated by averaging zip-code-level estimates from Aguilera et al. (2023).^33^ These estimates were derived using an ensemble machine learning approach, which achieved good performance across all sites (R^2^ = 0.83).^33^

Consistent with previous studies, we defined a smoke wave as at least 2 consecutive days when county-level wildfire smoke PM_2.5_ exceeded the 95^th^ percentile of the distribution of daily statewide average smoke PM_2.5_ concentrations over the study period (3.4 µg/m^3^).^2,34^ This definition was intended to capture air pollution episodes arising from one or more wildfire events.^2^ Successive smoke waves in the same county were combined and treated as a single event if the interval between the end of one smoke wave and the start of the next was 7 days or less (**Figure S1**). In sensitivity analyses, we replaced the primary smoke wave threshold with two alternative thresholds: (1) an absolute smoke PM_2.5_ threshold of 5 µg/m^3^ and (2) the 98^th^ percentile of the smoke PM_2.5_ concentration distribution (6.8 µg/m^3^).

### Estimation of excess acute care utilization during each smoke wave

We applied an ITS design combined with ML algorithms to estimate excess respiratory acute care encounters associated with each smoke wave in each county (**Figure 1**). For each smoke wave event, the goal was to estimate the counterfactual trend of respiratory acute care utilization that would have been expected in the absence of the smoke wave, and then compare this expected trend with the observed outcomes to estimate excess respiratory acute care encounters during the event following a three-step procedure.^29,30^

**Figure 1.**
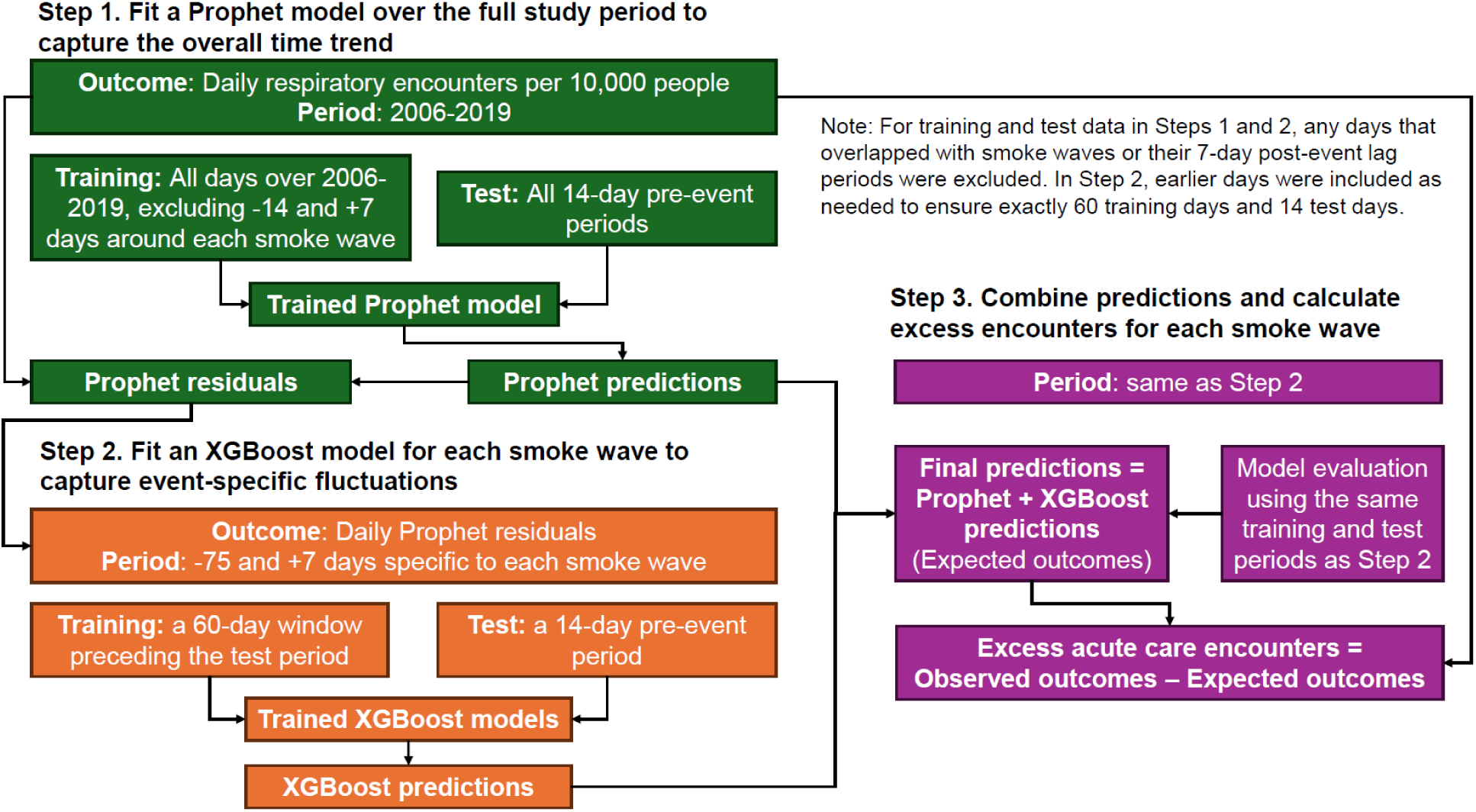
Flowchart of excess respiratory acute care utilization estimation. We used an interrupted time series design combined with Prophet-XGBoost models to estimate excess respiratory acute care encounters during each smoke wave in each county. This flowchart illustrates the main steps of this analytical framework.

In the first step, we fit a Prophet model for each county over the full study period to capture the overall temporal pattern in daily respiratory acute care encounters per 10,000 population, including long-term trends, recurring seasonal variation, day-of-the-week effects, and holiday effects. Prophet is an additive regression model for forecasting time-series data.^35^ To avoid contamination by smoke wave-related changes, we excluded all smoke wave days, along with the 14-day pre-event and 7-day post-event lag periods around all smoke waves, from model training. Based on the findings from previous studies, we assumed that the effects of smoke waves on respiratory acute care utilization were acute and lasted no more than 7 days.^29^ The 14-day periods immediately preceding each smoke wave were collectively used as the test period; any days within these test data that overlapped with smoke waves or their 7-day lag periods were excluded. The residuals of the fitted Prophet model represent short-term deviations not explained by the overall temporal trend.

In the second step, we fit an event-specific XGBoost model^36^ for each smoke wave to model the Prophet residuals and capture local fluctuations around the event. To balance model stability with temporal relevance to each event, we chose a 60-day training window preceding a 14-day pre-event test window. Any days that overlapped with other smoke waves and their lag periods were skipped when constructing both windows, and earlier days were included as needed to ensure exactly 60 training days and 14 test days. Predictors included calendar date and date-derived temporal features, holiday indicator, non-smoke PM_2.5_ concentration, maximum and minimum air temperatures, maximum and minimum relative humidity, and total precipitation (see data sources for these predictors in **Methods S1**). Hyperparameters were tuned over a space-filling grid of 30 candidate combinations using 5-fold rolling time-series cross-validation. The final hyperparameter combination was selected based on root mean square error (RMSE). The finalized XGBoost model was then fit and used to predict the residual component over the event-specific prediction window through 7 days after the smoke wave.

Finally, in the third step, we combined the Prophet predictions and the XGBoost-predicted residuals to obtain the final counterfactual encounters for each smoke wave event. For each smoke wave, model performance was evaluated using mean absolute scaled error (MASE), mean absolute error (MAE), and RMSE, calculated separately for the corresponding training and test periods. Excess respiratory acute care encounters were estimated as the difference between the observed outcomes and these expected outcomes during the smoke wave. To quantify uncertainty in the counterfactual predictions, we used a moving block bootstrap with a block length of 7 days and 50 bootstrap replicates to estimate the standard error of the predictions on each day and then constructed 95% confidence intervals (CIs) using a normal approximation.^37,38^

To summarize excess respiratory acute care encounters across smoke waves, we pooled the event-specific estimates to the state level using meta-regression models, with and without county fixed effects. We also pooled the estimates by county and by year to characterize spatial and temporal heterogeneity. In addition, we evaluated cumulative lagged effects for up to 7 days after the end of each smoke wave. Only smoke waves with MASE < 1 were included in this step, because values below 1 indicate that the model achieved, on average, smaller errors than the one-step naïve benchmark forecast.^39^

### Evaluation of factors that exacerbate the health impacts of smoke waves

To explore drivers of heterogeneity in excess respiratory acute care utilization across smoke waves, we examined potential effect modifiers in three categories: (1) smoke wave duration and timing; (2) smoke wave intensity; and (3) climate and meteorological conditions during the smoke wave. Smoke wave duration and timing included duration (number of days), season (spring, summer, fall, and winter), whether one or more smoke waves had occurred in the previous 30 days, whether the smoke wave overlapped with U.S. federal holidays, and diurnal patterns of smoke exposure (day-peak, night-peak, or homogeneous or tied) (see details in **Methods S2**). Smoke wave intensity was represented by concentrations of smoke PM_2.5_ and other smoke-related air pollutants during the smoke wave, including nitrogen dioxide (NO_2_), smoke ozone, and smoke particulate matter with aerodynamic diameter ≤ 10 μm (PM_10_). Climate and meteorological conditions included drought (no drought, mild drought, or severe drought), Santa Ana winds (present or absent; only applicable to counties in southern California), precipitation (present or absent), and extreme heat (present or absent) during each smoke wave. For continuous variables, we categorized values into tertiles and compared T2 and T3 with T1 to allow for potential nonlinearity. Details on data sources and variable categorization are provided in **Methods S2**. We analyzed each potential effect modifier separately using a random-effect meta-regression with county fixed effects to adjust for time-invariant differences across counties, with excess respiratory acute care encounters per 10,000 population during each smoke wave as the dependent variable. In sensitivity analyses, we also excluded county fixed effects or adjusted for season in meta-regressions. R packages *tidymodels*, *modeltime*, and *mvmeta* were used for these analyses.^40–42^ The datasets and R scripts used in this study are publicly available at https://github.com/benmarhnia-lab/Smokewave-ITS.

## Results

A total of 18,271,754 respiratory acute care encounters in California (2006-2019) were included in this study. Over the study period, 1,318 smoke waves were identified across the 56 included counties (**Figure S1**). The spatial distribution of smoke waves and respiratory acute care utilization in California from 2006 to 2019 is shown in **Figure 2**. The number of smoke waves varied substantially across years (**Figure S2**).

**Figure 2.**
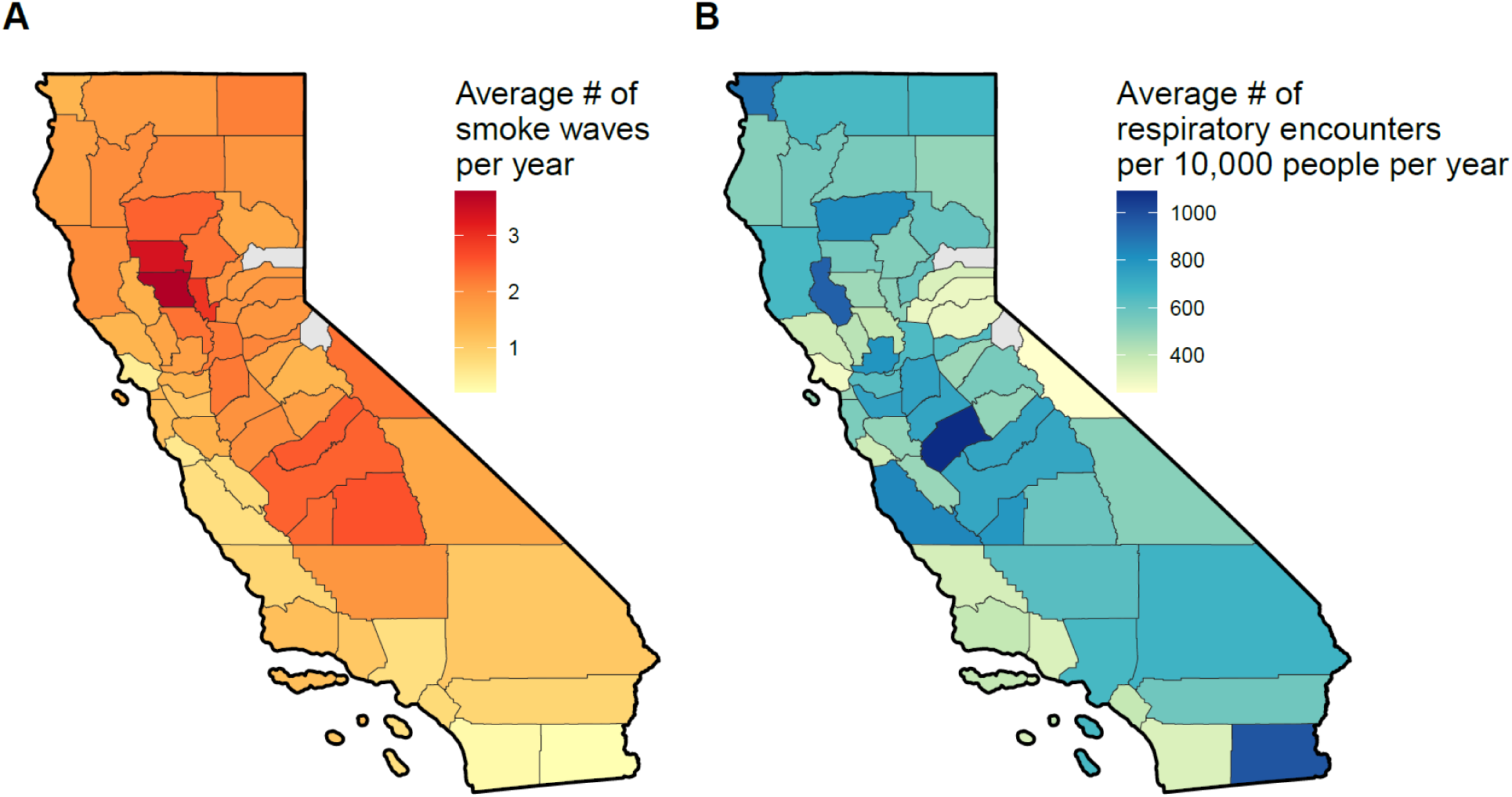
Spatial distribution of county-level smoke waves and respiratory acute care utilization in California, 2006-2019 (n = 1,318 smoke waves). This figure shows the county-level average annual number of smoke waves (A) and respiratory acute care encounters per 10,000 people (B) in California. The two gray counties (Alpine and Sierra) were excluded from this study because their populations were less than 5,000.

Using Prophet-XGBoost models with temporal features and additional predictors (**Table S1**), for each smoke wave, we estimated the expected trend in respiratory acute care utilization under the counterfactual scenario in which smoke waves did not occur. The models showed good performance across all counties, with mean training and test MASE values of 0.44 and 0.47, respectively (**Table S2**). **Figures 3A-3F** illustrate six selected examples, with one of the most intense events selected from each geographical region defined by the California Department of Fish and Wildlife.^43^ For example, during the smoke wave in San Diego County from October 21 to 29, 2007, 9.92 respiratory acute care encounters per 10,000 people were observed, whereas the expected number under the counterfactual scenario was 6.88 per 10,000 people (95% CI: 5.43, 8.33), indicating 3.04 excess encounters per 10,000 people (95% CI: 1.59, 4.49) attributable to the smoke wave (**Figure 3E**).

**Figure 3.**
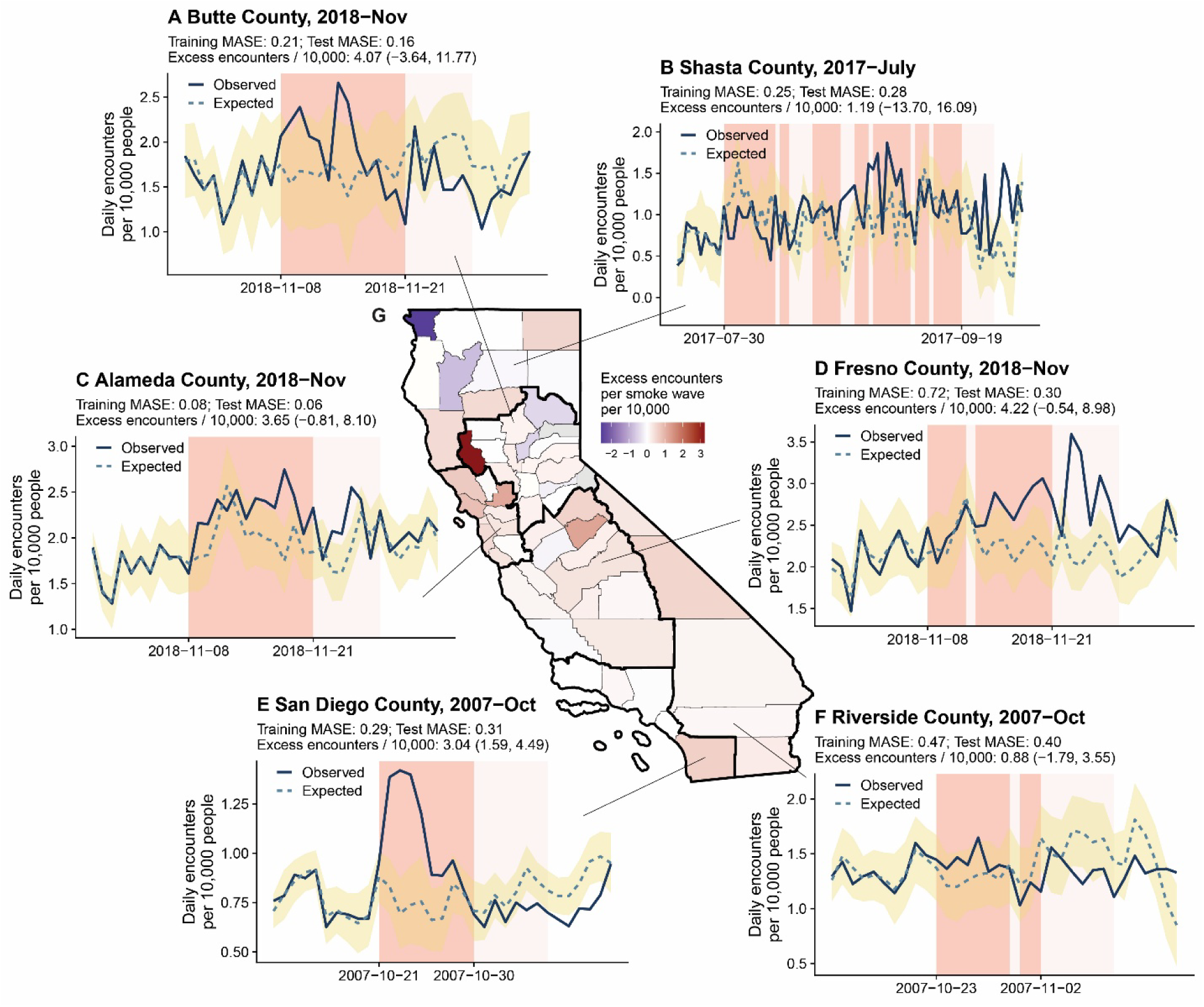
Observed and expected daily respiratory acute care utilization before, during, and after smoke waves, and pooled excess utilization per smoke wave in each county. Panels A-F illustrate six example smoke waves, one from each California region, and show the observed (solid line) and expected (dashed line) daily respiratory acute care encounters per 10,000 people before, during, and after each smoke wave. The dark pink background indicates days on which smoke PM_2.5_ concentrations exceeded the smoke wave threshold, and the light pink background indicates up to 7 subsequent days. The light yellow shaded area indicates the 95% confidence intervals of the expected values. The map in Panel G shows the pooled excess respiratory acute care encounters per 10,000 people per smoke wave in each county (n = 1,249 smoke waves). MASE: mean absolute scaled error.

When all smoke waves were pooled together, the mean excess number of encounters was 0.20 per 10,000 people per smoke wave event (95% CI: 0.12, 0.28) without county fixed effects and 0.51 per 10,000 people per smoke wave (95% CI: -0.10, 1.13) with county fixed effects. When higher smoke PM_2.5_ concentration thresholds were used to identify smoke waves, the estimated excess utilization increased, which likely reflected the greater severity of the identified smoke waves (**Figure S3**). When days immediately after each smoke wave were included, the excess estimates increased with longer post-event windows but stabilized within 6-7 lag days, suggesting lagged effects lasting up to about one week (**Figure S3**).

To explore heterogeneity across smoke waves, we pooled the event-specific excess acute care utilization estimates by county (**Figure 3G**) and by year (**Figure 4**). We observed substantial spatial heterogeneity in the pooled excess respiratory acute care encounters per smoke wave, with positive estimates in most counties and a few negative estimates scattered across Northern California. Substantial year-to-year variation was also observed. Positive excess estimates were observed in several years, whereas some negative estimates were observed in years such as 2010, with confidence intervals overlapping zero, likely due to the small number of smoke waves in those years (**Figure 4**). In the model with county fixed effects (**Figure S4**), the point estimates were generally similar. The variability of the pooled yearly estimates likely reflects the large variation in the number of smoke waves across years, but it may also indicate the presence of effect modifiers underlying the heterogeneity.

**Figure 4.**
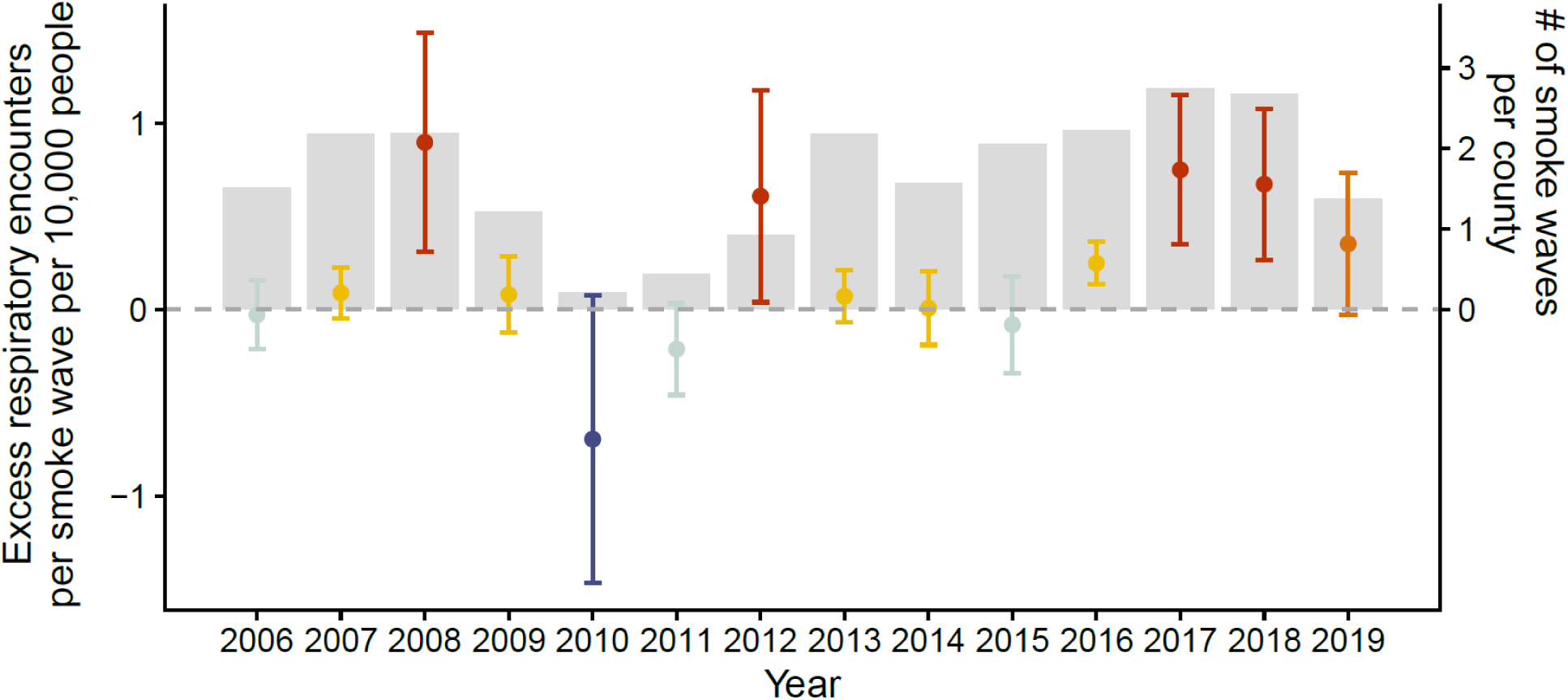
Pooled excess respiratory acute care encounters per smoke wave by year (n = 1,249 smoke waves). This figure shows the pooled excess respiratory acute care encounters per 10,000 people per smoke wave for each year, without county fixed effects in the meta-analysis. The error bars indicate 95% confidence intervals. Color indicates the magnitude and direction of the point estimate, with darker red representing more positive estimates and darker blue representing more negative estimates. The light gray bars show the average number of smoke waves per county each year.

**Figure 5** shows how excess respiratory acute care encounters changed with smoke wave duration and timing, intensity, and co-occurring climate and meteorological conditions, with county fixed effects included in the meta-regressions. Excess encounters were higher for smoke waves with longer duration (T3 [median: 13 days] vs. T1 [median: 2 days]). By season, smoke waves that occurred in winter had larger excess estimates than those in spring. Smoke waves that overlapped with holidays and those preceded by at least one smoke wave in the previous 30 days were associated with higher excess acute care utilization. No meaningful effect modification by diurnal pattern was observed. For intensity, higher smoke PM_2.5_, PM_10_ and NO_2_ levels during smoke waves were associated with larger excess respiratory acute care encounters, especially when comparing the highest with the lowest tertile. For climate and meteorological conditions, excess encounters were generally higher during smoke waves occurring under drought conditions, especially severe drought, and were elevated when smoke waves co-occurred with precipitation or extreme heat. Among smoke waves in southern California, the difference by Santa Ana wind occurrence was small. Results were generally consistent in models without county fixed effects and in models adjusted for season (**Figure S5 & S6**). The description of each tertile and the numbers of smoke waves in each category are provided in **Table S3**.

**Figure 5.**
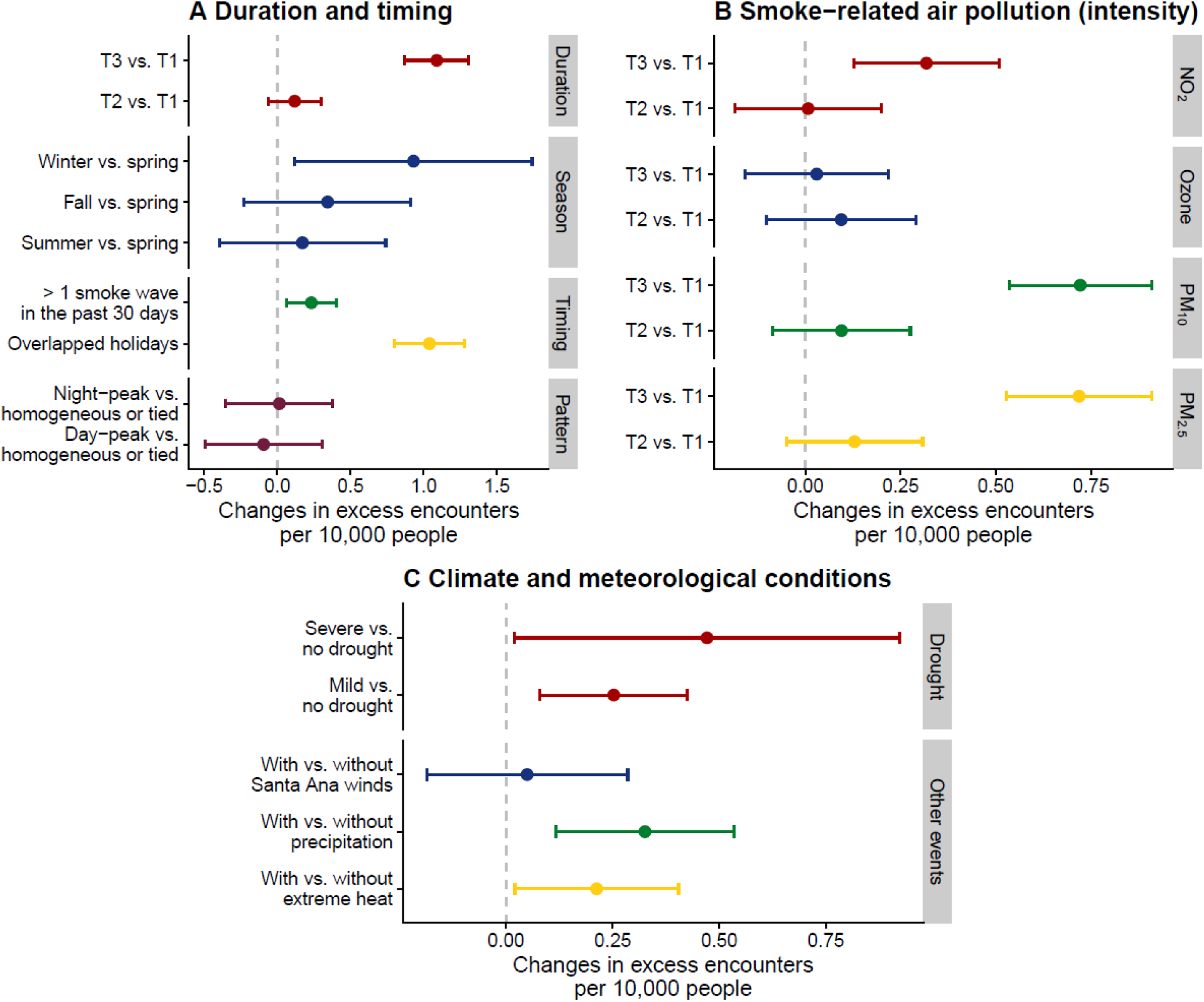
Effect modification by smoke wave duration, timing, intensity, and co-occurring meteorological conditions (n = 1,249 smoke waves). This figure shows how excess respiratory acute care encounters changed with smoke wave duration and timing (A), levels of smoke related air pollutants during smoke waves (B), and co-occurring climate and meteorological conditions (C), with county fixed effects included in the meta-regressions. T1, T2, and T3 denote tertiles. Santa Ana winds were only examined among smoke waves in southern California (n = 74).

## Discussion

This study presents the most comprehensive evaluation of spatiotemporal heterogeneity in respiratory health impacts across smoke waves using a novel causal inference framework. We found that greater excess respiratory acute care utilization was associated with smoke waves characterized by longer duration, higher intensity, winter occurrence, occurrence within 30 days of a prior smoke wave, overlap with holidays, drought conditions, and co-occurrence with precipitation or extreme heat.

The role of intensity in smoke wave-related respiratory disease burden is expected and consistent with previous studies.^16,17^ For example, a study in the western U.S. found that the association between exposure to smoke wave days and respiratory hospital admissions showed an increasing trend as smoke PM_2.5_ concentration increases.^16^ Our study suggests that, in addition to smoke PM_2.5_, higher concentrations of other smoke related air pollutants during the same event, such as NO_2_ and PM_10_, were also associated with greater excess respiratory acute care utilization.

The higher excess burden associated with winter smoke waves and those overlapping holidays may reflect differences in public preparedness, human behavior, and underlying respiratory vulnerability. In addition, cold, dry air can weaken airway defense mechanisms and facilitate respiratory virus transmission,^44^ potentially increasing susceptibility to smoke-related respiratory irritation. Similarly, smoke waves overlapping with holidays may coincide with reduced staffing or delayed response capacity in public agencies and health systems, as well as changes in population behavior, such as travel or increased outdoor activities, that could increase smoke exposure or alter care-seeking patterns.^45^ Although plausible, these conjectures remain speculative, and the underlying mechanisms warrant further investigation.

The higher excess respiratory acute care utilization observed during smoke waves preceded by another smoke wave within the previous 30 days suggests that, although consecutive events separated by ≤ 7 days intervals were combined into a single event, an 8-to-30-day interval between smoke waves may still exacerbate respiratory health impacts. This result aligns with previous occupational studies of firefighters indicating that repeated smoke exposure over a wildfire season may worsen airway inflammation and impair pulmonary function.^46,47^ This finding is particularly concerning because intervals between repeated smoke waves have shortened in California over the past two decades and may continue to shorten as wildfire smoke events become more frequent.^48^

Regarding co-occurring meteorological conditions, our findings align with the growing public health concern about compound climate hazards.^24^ Consistent with previous studies in California,^25–27^ we found that smoke waves co-occurring with extreme heat were associated with higher excess respiratory acute care utilization. Our study extends the existing literature by providing evidence that, beyond extreme heat, co-occurring drought conditions and precipitation may also exacerbate the respiratory impacts of smoke waves. Drought may amplify respiratory impacts by promoting drier fuels, higher smoke-related PM_2.5_ concentrations, and additional particle exposures such as dust.^49,50^ Precipitation may, however, have the potential to negatively impact respiratory symptoms by influencing concentrations, dispersion, or allergenicity of respiratory irritants or allergens.^51^

Existing wildfire smoke guidance is largely organized around current or forecasted air quality conditions,^52,53^ but the substantial heterogeneity observed in our study underscores the importance of treating smoke waves as discrete events with distinct health profiles. In addition, the findings also suggest unique opportunities for novel mitigation or adaptation guidance.^47,54^

A few limitations should be noted. First, the identification of smoke waves depends on the source of smoke PM_2.5_ data, threshold choices, spatial aggregation, minimum duration, and rules for merging nearby smoke episodes; alternative definitions could yield different event sets and health impact estimates. Our main results remained robust when applying alternative smoke wave thresholds in sensitivity analyses. Second, the ML-integrated ITS approach relies on the ability of the algorithms and pre-event data to predict counterfactual trends during the smoke wave period. In our analysis, we excluded 67 smoke waves (5%) with poor model performance from the final stage; however, these excluded events showed no apparent spatial or temporal clustering, suggesting that the exclusions were unlikely to introduce systematic bias.

## Conclusions

As wildfire activity is projected to intensify, incorporating event characteristics and compound meteorological conditions into wildfire smoke preparedness and response planning may help identify especially harmful smoke waves and better protect respiratory health. By moving beyond average smoke-health associations, these findings provide event-specific assessments of respiratory health impacts and can inform more targeted wildfire smoke preparedness, risk communication, and public health response.

## Funding

This work was supported by the California Air Resources Board (CARB) under award number 23RD005 and the NIH Climate Opportunities for Health (ECOHEALTH) Center (1P20MD019994-01) via Secondary Analysis Pilot Program.

## Author contributions

Y.M. conducted formal analyses and drafted the manuscript. A.K.D. contributed to the study design and editing of the manuscript. R.A., A.G., and A.W. contributed to data collection and edited the manuscript. N.L., P.S., C.G.J., M.J., R.C., N.T., and M.E.M. contributed to result interpretation and edited the manuscript. T.B. contributed to the study design, conceived of and supervised the conduct of this study, and edited the manuscript.

## Conflicts of interest

The authors declare they have no conflicts of interest related to this work to disclose.

## Data availability statement

The datasets and R scripts used in this study are publicly available at https://github.com/benmarhnia-lab/Smokewave-ITS.

## Ethical approval statement

This study was approved by the California Health and Human Services Agency’s Committee for the Protection of Human Subjects (project number: 2021-116).

## Notes

### Competing Interest Statement

The authors have declared no competing interest.

### Author Declarations

This study was approved by the California Health and Human Services Agency Committee for the Protection of Human Subjects (project number: 2021-116).

