## Supplementary material for "Heterogeneity in excess respiratory acute care utilization during wildfire smoke waves in California"

**Methods S1. Data sources for machine learning model predictors**

In addition to calendar date, holiday indicators, and other temporal features derived from the date variable, we included the following variables as predictors in the machine learning model. Their corresponding data sources and aggregation methods are also provided in the table.

| Variable | Data source | Aggregation |
|---|---|---|
| Non-smoke $PM_{2.5}$ concentration | Aguilera et al. (2023)[1] | County-level average of ZIP-code-level concentrations |
| Maximum and minimum air temperatures | Gridded Surface Meteorological dataset (GridMet)[2] | Area-weighted county-level average of 4-$km^2$ grid values |
| Maximum and minimum relative humidity | GridMet[2] | Area-weighted county-level average of 4-$km^2$ grid values |
| Total precipitation | GridMet[2] | Area-weighted county-level average of 4-$km^2$ grid values |

**Methods S2. Data sources and categorization for potential effect modifiers**

This table lists the effect modifiers examined in this study, along with their data sources and the corresponding categorization or definition of each variable.

| Effect modifier | Definition | Data source | Categorization |
|---|---|---|---|
| ***Smoke-wave duration and timing*** | | | |
| Duration | Number of days with smoke $PM_{2.5}$ concentration above the smoke-wave threshold | / | Tertiles across all smoke waves (T1*, T2, T3) |
| Season | Season of occurrence, defined by the first day of the smoke wave | / | Spring (March to May)*; Summer (June to August); Fall (September to November); Winter (December to February) |
| Prior smoke waves within 30 days | Whether one or more smoke waves occurred within 30 days before the start of the smoke wave | / | Yes vs. No* |
| Holiday overlap | Whether the smoke wave overlapped with one or more holidays | U.S. federal public holidays | Yes vs. No* |
| Diurnal pattern | Classification of smoke waves according to within-day variability in hourly $PM_{2.5}$ concentrations and the timing of the daily peak $PM_{2.5}$ concentration. | Hourly $PM_{2.5}$ concentrations from EPA Air Quality System monitoring stations; county-level average across all stations within each county | Homogeneous or tied*; Day-peak; Night-peak<br><br>Please see table notes for more details.† |
| ***Smoke-wave intensity*** | | | |
| Smoke $PM_{2.5}$ | Average smoke $PM_{2.5}$ concentration during the smoke wave | Aguilera et al. (2023);[1] county-level average of ZIP-code-level concentrations | County-specific tertiles (T1*, T2, T3) |
| Smoke $PM_{10}$ | Average smoke $PM_{10}$ concentration during the smoke wave | Harvard Dataverse;[3,4] county-level average of ZIP-code-level concentrations | County-specific tertiles (T1*, T2, T3) |
| Smoke ozone | Average maximum 8-hour ozone concentration during the smoke wave | Harvard Dataverse;[3,4] county-level average of ZIP-code-level concentrations | County-specific tertiles (T1*, T2, T3) |
| Nitrogen dioxide ($NO_2$) | Average maximum 1-hour $NO_2$ concentration during the smoke wave | Harvard Dataverse;[5,6] county-level average of ZIP-code-level concentrations | County-specific tertiles (T1*, T2, T3) |
| ***Meteorological conditions*** | | | |

| | | | |
|---|---|---|---|
| Drought | Standardized precipitation evapotranspiration index (SPEI) at 1-month scale[7] | Global SPEI database;[8] area-weighted county-level average of 0.5-degree grid values | No drought (SPEI ≥ 0.5)*; Mild drought (-1.3 < SPEI < - 0.5); Severe drought (SPEI ≤ -1.3) |
| Santa Ana winds | During the smoke wave, at least one day with Santa Ana wind speed exceeding 0. Only applicable to southern California.‡ | ERA5;[9] area-weighted county-level average of 0.23-degree grid values | Present vs. Absent* |
| Precipitation | During the smoke wave, at least one day with precipitation exceeding 0. | GridMet;[2] area-weighted county-level average of 4-$km^2$ grid values | Present vs. Absent* |
| Extreme heat | During the smoke wave, at least one day with heat index (HI) exceeding the county-specific $98^{th}$ percentile of HI values during climate reference period (1981-2010) | GridMet;[2] area-weighted county-level average of 4-$km^2$ grid values | Present vs. Absent* |

*The category used as the reference group.

†For each smoke wave day, diurnal $PM_{2.5}$ patterns were classified based on within-day variability (amplitude) and the timing of the diurnal peak concentration (peak hour). Days with amplitudes below the $40^{th}$ percentile were classified as homogeneous, whereas days with higher amplitudes were classified as either night/early-morning peak (peak before 09:00 or after 20:00) or daytime peak (peak after 09:00 and before 20:00). Smoke wave types were then assigned at the event level by identifying the longest sequence of at least two consecutive days belonging to the same class within each smoke wave. The dominant sequence determined whether the event was classified as homogeneous, night-peak, or day-peak; ties were assigned when no single class had a uniquely longest run. Hourly $PM_{2.5}$ data were unavailable for 9 of the 56 studied California counties (Amador, El Dorado, Lake, Lassen, Modoc, Plumas, Shasta, Tuolumne, and Yuba) and these counties were excluded when examining the effect modification by diurnal pattern.

‡Santa Ana winds were only applicable to and examined among smoke waves in the following southern California counties: Santa Barbara, Ventura, Los Angeles, San Bernardino, Riverside, Orange, and San Diego.

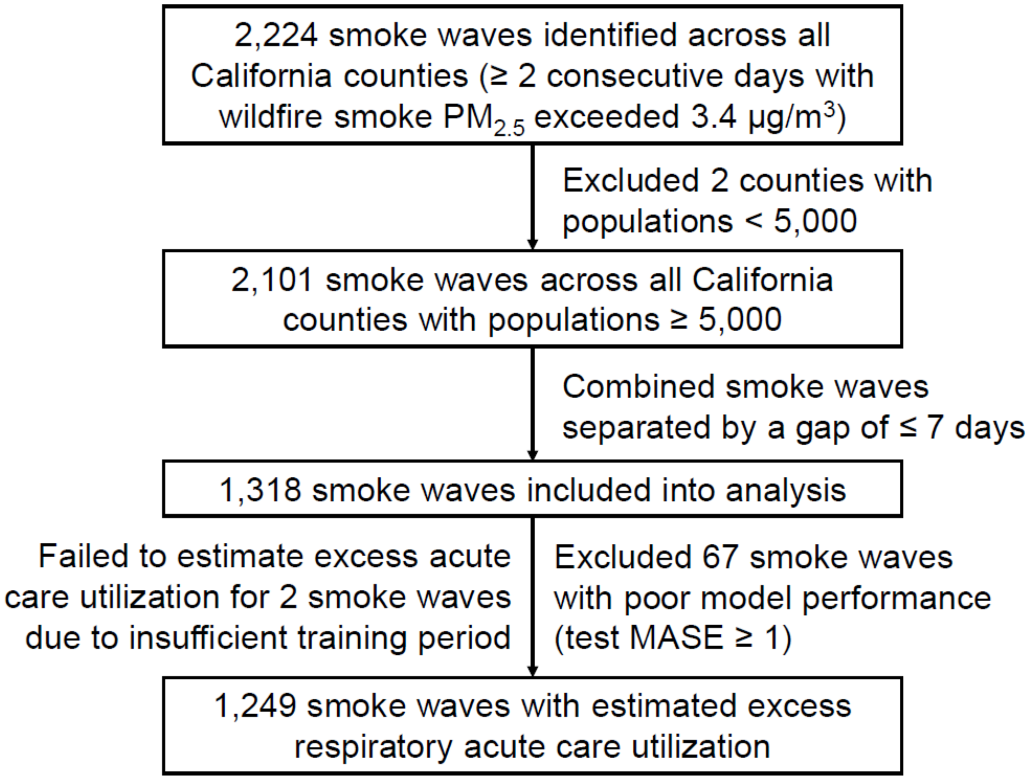


**Figure S1. Flowchart of smoke-wave counts at each step.**

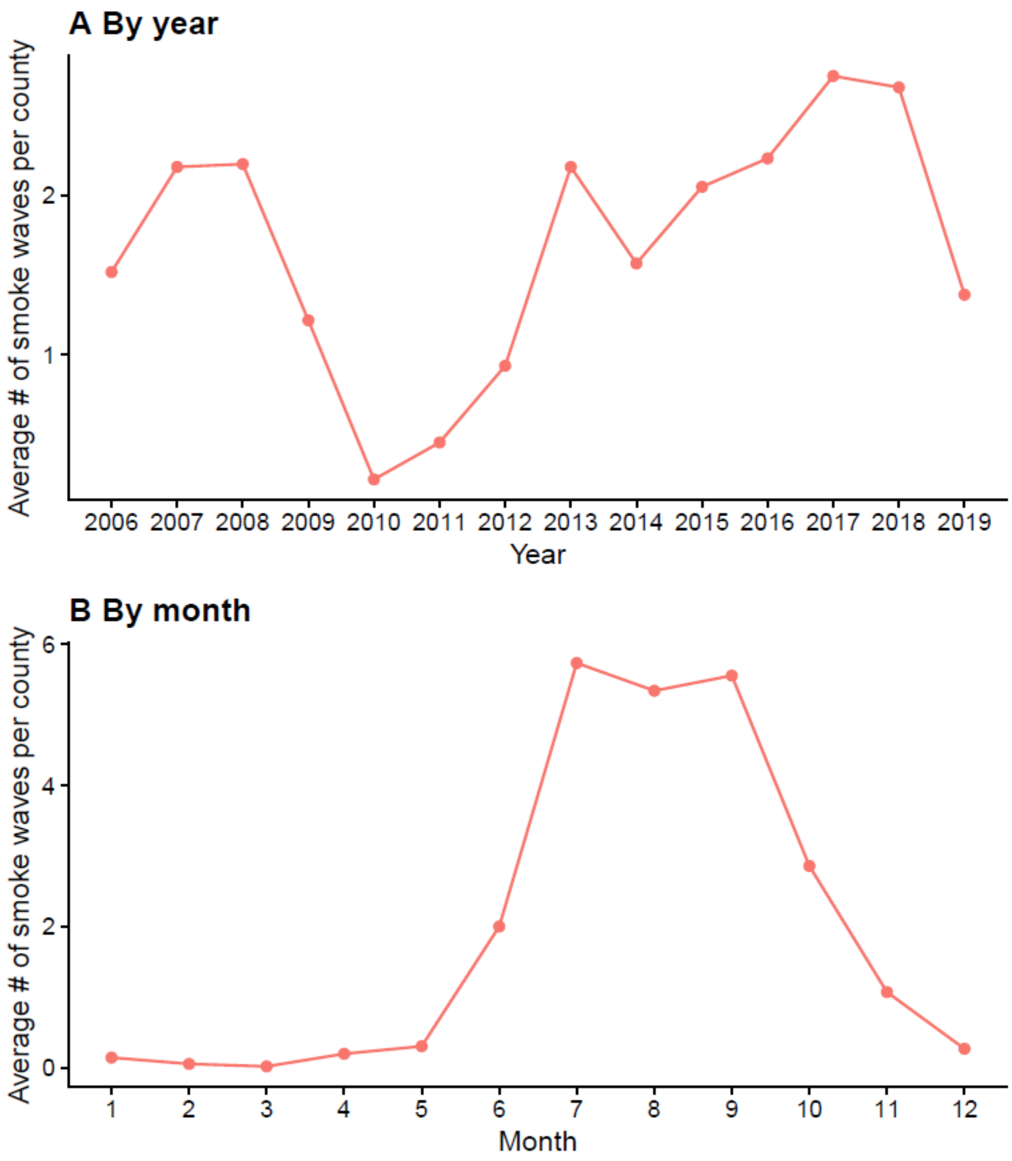


**Figure S2. County-level average number of smoke waves per year (A) and per month (B) in California, 2006-2019 (n = 1,318 smoke waves).**

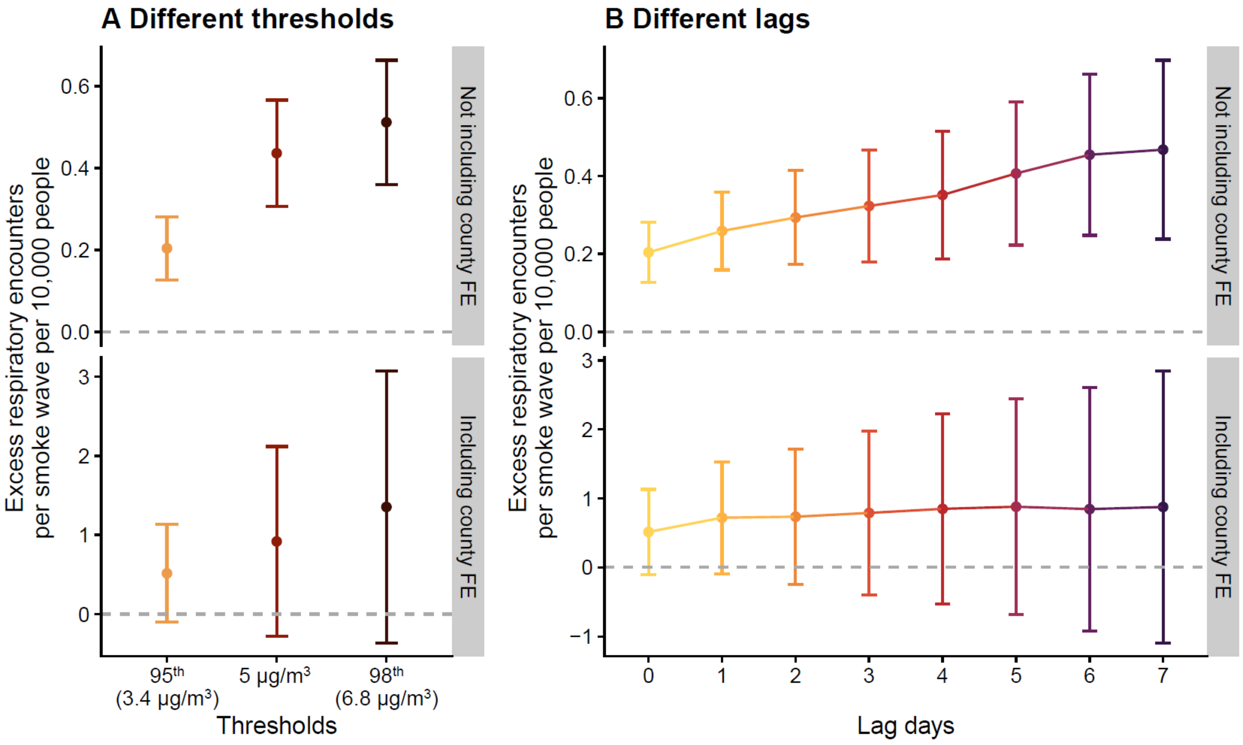


**Figure S3. Pooled state-level excess respiratory acute care encounters per smoke wave.**

Panel A shows the pooled excess respiratory acute care encounters per 10,000 people per smoke wave across all included counties, 2006-2019, when using the 95th percentile of the smoke $PM_{2.5}$ concentration distribution (3.4 μg/m$^3$, the main analysis; identified 1,249 smoke waves), an absolute threshold of 5 μg/m$^3$ (identified 904 smoke waves) and the 98th percentile of the distribution (6.8 μg/m$^3$; identified 667 smoke waves). Panel B shows the pooled state-level estimates when we included up to 7 lag days.

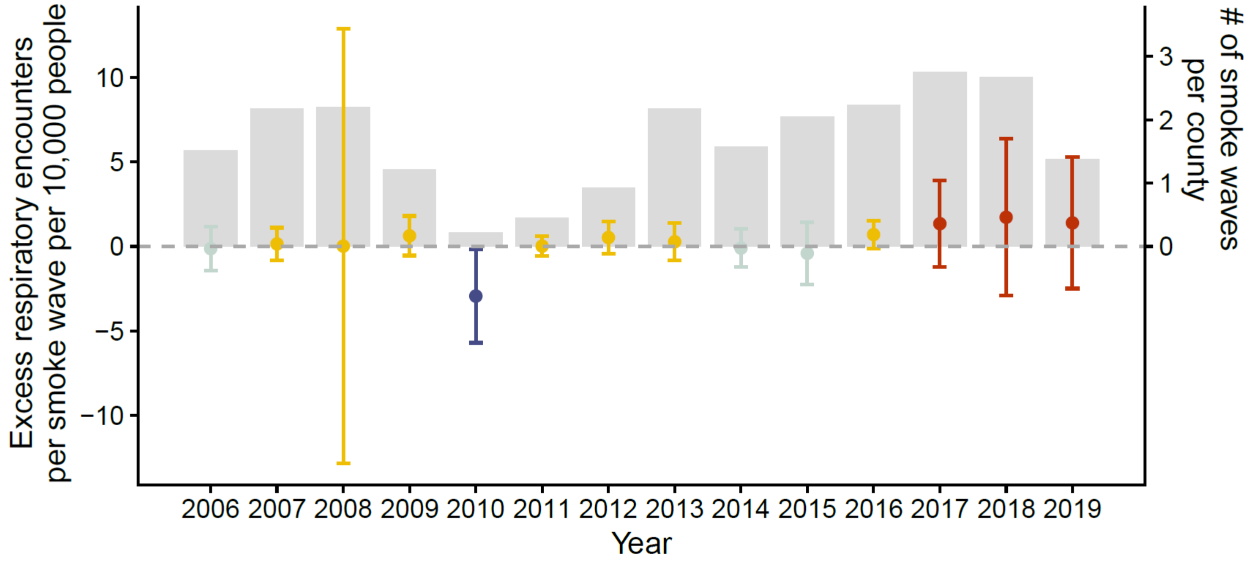


**Figure S4. Pooled excess respiratory acute care encounters per smoke wave by year, including county fixed effects (n = 1,249 smoke waves).**

This figure shows the pooled excess respiratory acute care encounters per 10,000 people per smoke wave for each year, with county fixed effects in the meta-analysis. The error bars indicate 95% confidence intervals. Color indicates the magnitude and direction of the point estimate, with darker red representing more positive estimates and darker blue representing more negative estimates. The light gray bars show the average number of smoke waves per county each year.

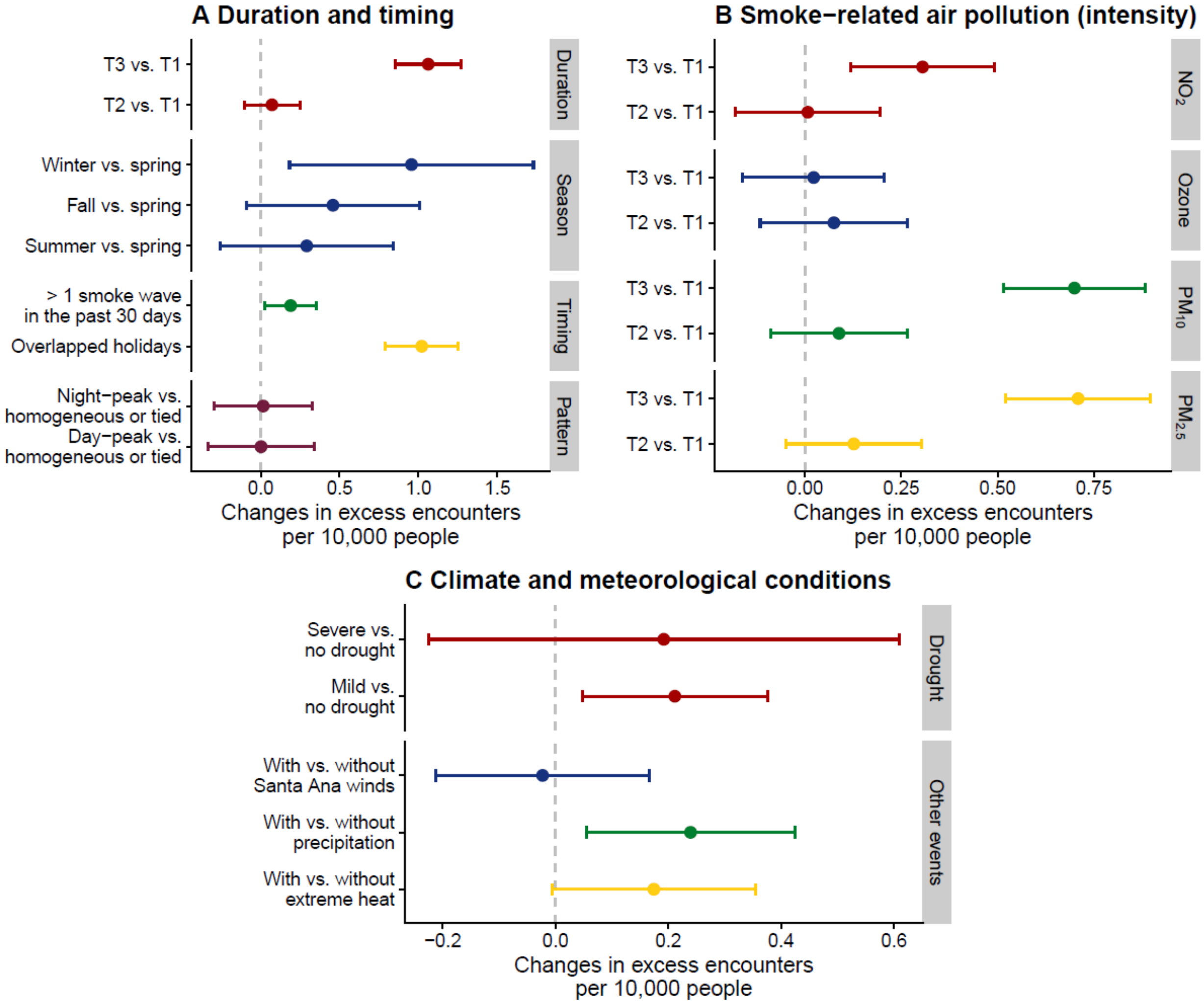


**Figure S5. Effect modification by smoke wave duration, timing, intensity, and co-occurring meteorological conditions, without county fixed effects in meta-regressions (n = 1,249 smoke waves).**

This figure shows how excess respiratory acute care encounters changed with smoke-wave duration and timing (A), levels of smoke related air pollutants during smoke waves (B), and co-occurring climate and meteorological conditions (C), without county fixed effects included in the meta-regressions. Santa Ana winds were only examined among smoke waves in southern California (n = 74).

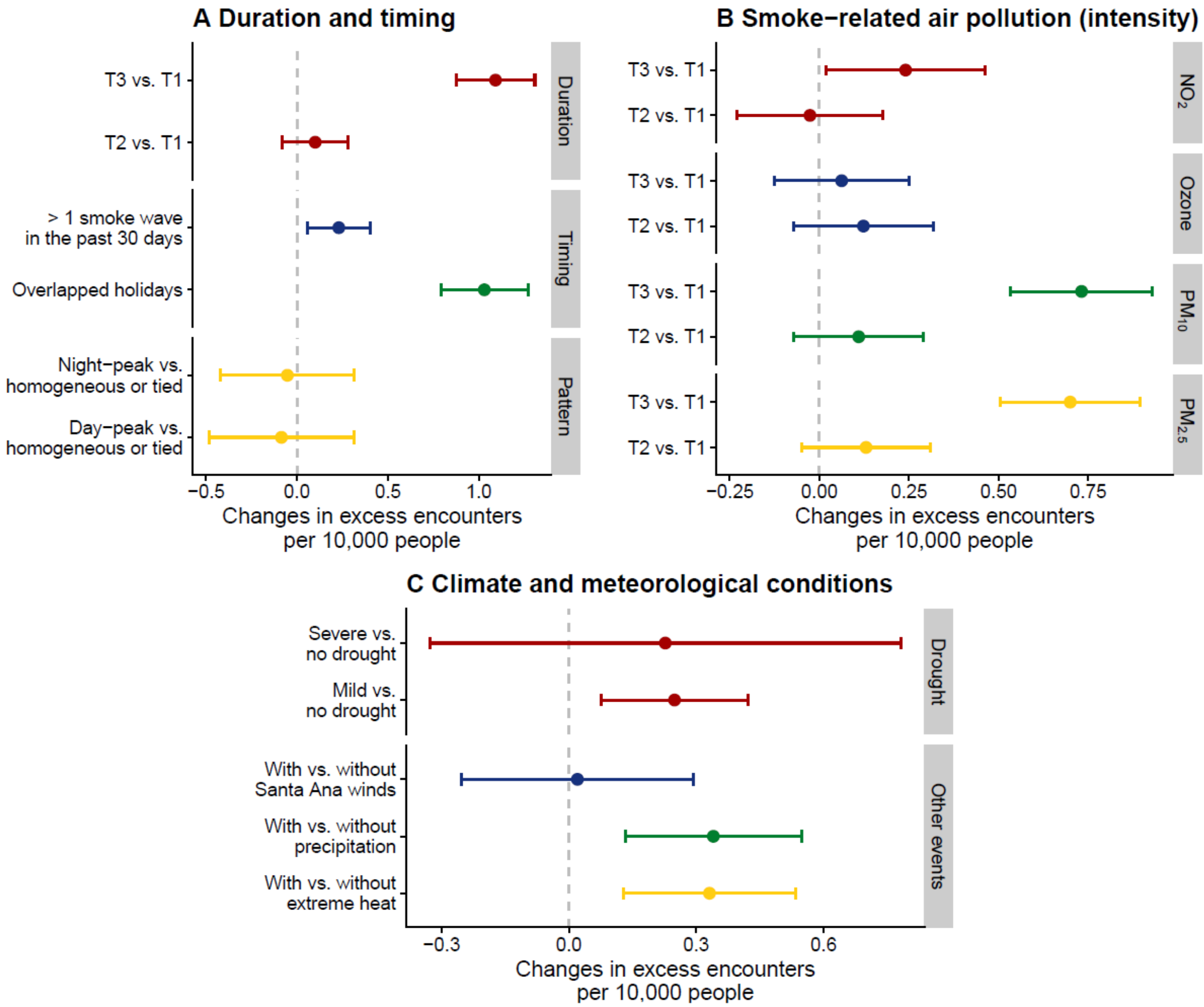


**Figure S6. Effect modification by smoke wave duration, timing, intensity, and co-occurring meteorological conditions, adjusted for season in meta-regressions (n = 1,249 smoke waves).**

This figure shows how excess respiratory acute care encounters changed with smoke-wave duration and timing (A), levels of smoke related air pollutants during smoke waves (B), and co-occurring climate and meteorological conditions (C), adjusting for season in the meta-regressions. Santa Ana winds were only examined among smoke waves in southern California (n = 74).

**Table S1. Descriptive statistics for daily county-level respiratory acute care encounters and machine learning model predictors in California, 2006-2019 (n = 286,328 county-days)**

| | Mean (SD) | Min | P5 | P25 | Median | P75 | P95 | Max |
|---|---|---|---|---|---|---|---|---|
| Respiratory acute care encounters per 10,000 people | 1.56 (1.00) | 0.00 | 0.39 | 0.88 | 1.35 | 2.01 | 3.46 | 17.55 |
| Non-smoke $PM_{2.5}$ (μg/m$^3$) | 7.62 (4.55) | 0.13 | 2.87 | 4.84 | 6.65 | 9.12 | 15.37 | 75.50 |
| Maximum temperature (˚C) | 22.28 (8.36) | -9.52 | 9.25 | 15.96 | 21.94 | 28.51 | 36.12 | 48.70 |
| Minimum temperature (˚C) | 8.40 (6.09) | -25.16 | -1.59 | 4.33 | 8.57 | 12.51 | 18.07 | 33.36 |
| Maximum relative humidity (%) | 79.03 (18.41) | 6.88 | 43.24 | 66.70 | 83.87 | 94.70 | 100.00 | 100.00 |
| Minimum relative humidity (%) | 35.09 (17.22) | 1.00 | 10.64 | 21.45 | 33.36 | 46.64 | 66.27 | 100.00 |
| Total precipitation (mm) | 1.69 (6.31) | 0.00 | 0.00 | 0.00 | 0.00 | 0.03 | 10.80 | 188.17 |

SD: standard deviation.

**Table S2. Model performance (n = 1,249 smoke waves)**

| County | # of smoke waves | Training | | | Test | | |
|---|---|---|---|---|---|---|---|
| | | MASE | MAE | RMSE | MASE | MAE | RMSE |
| Alameda | 15 | 0.45 | 0.07 | 0.09 | 0.40 | 0.06 | 0.08 |
| Amador | 29 | 0.42 | 0.28 | 0.36 | 0.49 | 0.30 | 0.38 |
| Butte | 27 | 0.54 | 0.16 | 0.20 | 0.59 | 0.16 | 0.20 |
| Calaveras | 23 | 0.44 | 0.24 | 0.32 | 0.50 | 0.24 | 0.31 |
| Colusa | 53 | 0.47 | 0.38 | 0.48 | 0.51 | 0.37 | 0.47 |
| Contra Costa | 19 | 0.36 | 0.06 | 0.07 | 0.36 | 0.06 | 0.08 |
| Del Norte | 19 | 0.33 | 0.39 | 0.51 | 0.36 | 0.37 | 0.47 |
| El Dorado | 26 | 0.54 | 0.12 | 0.15 | 0.60 | 0.10 | 0.13 |
| Fresno | 31 | 0.48 | 0.10 | 0.12 | 0.45 | 0.09 | 0.12 |
| Glenn | 46 | 0.35 | 0.29 | 0.37 | 0.40 | 0.30 | 0.38 |
| Humboldt | 22 | 0.44 | 0.17 | 0.21 | 0.45 | 0.17 | 0.20 |
| Imperial | 3 | 0.47 | 0.20 | 0.24 | 0.46 | 0.25 | 0.30 |
| Inyo | 22 | 0.42 | 0.42 | 0.54 | 0.49 | 0.42 | 0.51 |
| Kern | 25 | 0.33 | 0.06 | 0.07 | 0.33 | 0.06 | 0.07 |
| Kings | 31 | 0.44 | 0.22 | 0.28 | 0.45 | 0.21 | 0.26 |
| Lake | 19 | 0.40 | 0.29 | 0.37 | 0.44 | 0.32 | 0.40 |
| Lassen | 23 | 0.48 | 0.36 | 0.45 | 0.50 | 0.33 | 0.41 |
| Los Angeles | 8 | 0.21 | 0.02 | 0.02 | 0.27 | 0.03 | 0.03 |
| Madera | 35 | 0.43 | 0.19 | 0.24 | 0.47 | 0.18 | 0.22 |
| Marin | 5 | 0.60 | 0.09 | 0.12 | 0.56 | 0.11 | 0.14 |
| Mariposa | 24 | 0.38 | 0.36 | 0.47 | 0.47 | 0.34 | 0.43 |
| Mendocino | 28 | 0.54 | 0.27 | 0.34 | 0.56 | 0.26 | 0.32 |
| Merced | 27 | 0.32 | 0.15 | 0.19 | 0.38 | 0.16 | 0.20 |
| Modoc | 29 | 0.27 | 0.42 | 0.57 | 0.40 | 0.46 | 0.58 |
| Mono | 30 | 0.45 | 0.28 | 0.36 | 0.52 | 0.29 | 0.37 |
| Monterey | 9 | 0.45 | 0.15 | 0.19 | 0.47 | 0.17 | 0.21 |
| Napa | 22 | 0.52 | 0.18 | 0.23 | 0.56 | 0.17 | 0.20 |
| Nevada | 25 | 0.52 | 0.16 | 0.20 | 0.59 | 0.17 | 0.22 |
| Orange | 10 | 0.28 | 0.03 | 0.03 | 0.33 | 0.03 | 0.04 |
| Placer | 25 | 0.56 | 0.10 | 0.12 | 0.58 | 0.09 | 0.11 |
| Plumas | 22 | 0.40 | 0.38 | 0.49 | 0.44 | 0.40 | 0.50 |
| Riverside | 11 | 0.38 | 0.04 | 0.06 | 0.44 | 0.04 | 0.05 |
| Sacramento | 27 | 0.44 | 0.07 | 0.10 | 0.39 | 0.06 | 0.07 |
| San Benito | 11 | 0.30 | 0.15 | 0.19 | 0.39 | 0.18 | 0.21 |
| San Bernardino | 12 | 0.42 | 0.06 | 0.07 | 0.44 | 0.05 | 0.06 |
| San Diego | 4 | 0.38 | 0.03 | 0.04 | 0.36 | 0.03 | 0.03 |
| San Francisco | 18 | 0.52 | 0.10 | 0.12 | 0.50 | 0.09 | 0.11 |
| San Joaquin | 28 | 0.49 | 0.12 | 0.15 | 0.50 | 0.11 | 0.14 |
| San Luis Obispo | 10 | 0.45 | 0.10 | 0.12 | 0.46 | 0.09 | 0.11 |
| San Mateo | 17 | 0.61 | 0.11 | 0.14 | 0.56 | 0.10 | 0.13 |
| Santa Barbara | 16 | 0.55 | 0.11 | 0.13 | 0.58 | 0.11 | 0.13 |
| Santa Clara | 18 | 0.40 | 0.05 | 0.06 | 0.40 | 0.06 | 0.07 |
| Santa Cruz | 8 | 0.34 | 0.10 | 0.13 | 0.43 | 0.11 | 0.14 |
| Shasta | 27 | 0.47 | 0.14 | 0.18 | 0.53 | 0.15 | 0.19 |
| Siskiyou | 25 | 0.49 | 0.34 | 0.43 | 0.57 | 0.36 | 0.46 |
| Solano | 20 | 0.54 | 0.17 | 0.22 | 0.61 | 0.18 | 0.22 |
| Sonoma | 18 | 0.49 | 0.09 | 0.11 | 0.53 | 0.08 | 0.10 |
| Stanislaus | 24 | 0.51 | 0.11 | 0.14 | 0.50 | 0.11 | 0.13 |

| | | | | | | | |
|---|---|---|---|---|---|---|---|
| Sutter | 39 | 0.41 | 0.19 | 0.25 | 0.45 | 0.18 | 0.23 |
| Tehama | 33 | 0.45 | 0.27 | 0.34 | 0.46 | 0.27 | 0.34 |
| Trinity | 28 | 0.27 | 0.31 | 0.42 | 0.37 | 0.36 | 0.47 |
| Tulare | 34 | 0.46 | 0.11 | 0.13 | 0.46 | 0.11 | 0.14 |
| Tuolumne | 19 | 0.37 | 0.20 | 0.25 | 0.42 | 0.21 | 0.27 |
| Ventura | 13 | 0.51 | 0.06 | 0.08 | 0.50 | 0.05 | 0.07 |
| Yolo | 31 | 0.48 | 0.14 | 0.18 | 0.49 | 0.13 | 0.16 |
| Yuba | 26 | 0.31 | 0.17 | 0.22 | 0.40 | 0.18 | 0.23 |
| **All counties** | **1,249** | **0.44** | **0.20** | **0.26** | **0.47** | **0.20** | **0.25** |

MASE: mean absolute scaled error

MAE: mean absolute error

RMSE: root mean squared error

**Table S3. Descriptive statistics for examined effect modifiers (n = 1,249 smoke waves)**

| | Median (IQR) or n (%) |
|---|---|
| Duration (days) | |
| T1 | 2 (1) |
| T2 | 5 (1) |
| T3 | 13 (18) |
| Mean smoke $PM_{2.5}$ (μg/m$^3$) | |
| T1 | 4.88 (1.30) |
| T2 | 7.12 (1.89) |
| T3 | 12.85 (9.70) |
| Mean maximum 1-hour $NO_2$ (ppb) | |
| T1 | 10.82 (5.83) |
| T2 | 15.68 (8.59) |
| T3 | 22.06 (11.46) |
| Mean smoke maximum 8-hour ozone (ppb) | |
| T1 | 1.84 (3.01) |
| T2 | 5.19 (4.81) |
| T3 | 9.06 (7.38) |
| Mean smoke $PM_{10}$ (μg/m$^3$) | |
| T1 | 6.58 (6.06) |
| T2 | 12.68 (7.59) |
| T3 | 23.26 (16.76) |
| Season | |
| Spring | 28 (2.2%) |
| Summer | 687 (55.0%) |
| Fall | 510 (40.8%) |
| Winter | 24 (1.9%) |
| Overlapped holidays | |
| Yes | 241 (19.3%) |
| No | 1,008 (80.7%) |
| # of smoke waves over the past 30 days | |
| 0 | 440 (35.2%) |
| 1 or 2 | 809 (64.8%) |
| Diurnal pattern | |
| Day-peak | 122 (9.8%) |
| Night-peak | 158 (12.7%) |
| Homogeneous or tied | 337 (27.0%) |
| Not applicable | 632 (50.6%) |
| Drought (measured by SPEI-1) | |
| No drought (SPEI-1 ≥ -0.4) | 801 (64.1%) |
| Mild drought (-1.3 < SPEI-1 < -0.4) | 407 (32.6%) |
| Severe drought (SPEI-1 ≤ -1.3) | 41 (3.3%) |
| Santa Ana Winds (only applicable to Southern California) | |
| With Santa Ana Winds | 32 (2.6%) |
| Without Santa Ana Winds | 42 (3.4%) |
| Not applicable | 1,175 (94.1%) |
| Precipitation | |
| With precipitation | 355 (28.4%) |
| Without precipitation | 894 (71.6%) |
| Extreme heat | |
| With extreme heat | 353 (28.3%) |

| | |
|---|---|
| Without extreme heat | 896 (71.7%) |
